# An investigation into the wellbeing of optometry students

**DOI:** 10.1101/2022.03.17.22272543

**Authors:** Anne Vorster, Barbara M Hamlett, Janike le Roux, Janke Blount, Jane Knoesen, Lauren S Coetzee

**Affiliations:** Department of Optometry, University of the Free State, South Africa; Department of Optometry and Vision Sciences, University of Huddersfield, United Kingdom; Division of Health Sciences Education, University of the Free State, South Africa

**Keywords:** COVID-19, student wellbeing, optometry students, anxiety, depression

## Abstract

**Introduction:** Wellbeing is synonymous with positive mental health and impacts the efficacy of student learning. The wellbeing of optometry students is an understudied topic. The wellbeing of optometry students studying in a blended undergraduate course during the COVID-19 pandemic was also unknown. The aim of this study was to determine the status of optometry students’ wellbeing during COVID-19, by identifying their experiences of symptoms of anxiety and depression. Moreover, to determine to what extent students experience these symptoms and what specific factors influenced their wellbeing.

**Methodology:** Participants from four year groups completed online questionnaires. The response rate was 78.38% (n=87). Zung self-rating depression and anxiety scale questionnaires were used to determine whether students identified with a given list of symptoms commonly linked to anxiety or depression. Through open ended questions students’ wellbeing was further investigated.

**Results:** Participants experienced normal levels of anxiety symptoms and most participants experienced mild to moderate depression symptoms. Of concern is the severe depression symptoms identified in the third and fourth year student cohorts. Mental health, Academics, Lifestyle, Relationships and Sleep were main themes identified that had an influence on the students’ general wellbeing. Uncertainty and Physical Health themes were additional influences of wellbeing specifically related to COVID-19.

**Contribution:** This preliminary study into wellbeing of optometry students was undertaken in a unique timeframe, due to the COVID-19 pandemic. The results provide a platform to determine baseline wellbeing in the future student cohorts and the exploratory identification of factors causing stress and anxiety. The impact of COVID-19 on the wellbeing of students is evident.

**Conclusion:** Optometry students do experience symptoms of depression. COVID-19 has had considerable impact on their academic experience and their wellbeing.

## Background

Mental health and wellbeing have been topics of great concern recently. Ordinary people around the world endured many weeks of separation and isolation, with limited human engagement. Wellbeing is synonymous with positive mental health. Wellbeing has been defined as the combination of feeling positive emotions, such as joy and satisfaction, having a sense of purpose and realising one’s potential and the development of abilities, whilst coping with stressors, having some control over one’s life and engaging in positive relationships (1).

Wellbeing has been associated with success in both professional and personal lives of individuals, as well as more effective learning in students (1). Greater subjective well-being has been found to correlate with higher academic performance (2). Higher status of wellbeing has been linked to better physical health, higher life satisfaction and better work performance (1).

When environmental demands are more than an individual can effectively handle and could potentially cause physical and psychological problems, it is known as stress (3, 4). How students experience and handle stress may negatively influence their academic performance, cause depression and may lead to health problems, and even suicide (5, 6). Stress, anxiety, and depression are all factors contributing to the wellbeing of an individual and influence how people live and experience life. Studies have been conducted on the occurrence of stress and diseases caused by stress such as depression and anxiety experienced by undergraduate students (3, 6, 7). The transition to university life amongst first year students forces the development of social skills, responsibility and learning to cope without family and friends. Moreover, stress caused from expected academic performance as well as time pressures tests, required grades, higher standard of academic work and anxieties about future job opportunities (5, 8).

A correlation has been established between academic performance and the wellbeing of students (7). Medical students experience stress, anxiety and depression due to the academic and clinical challenges (3, 5, 6). Often health-profession students, put patients’ needs before their own, leading to an underreporting of mental health conditions in health-profession students (8). Wellbeing of health-profession students has also been found to be negatively impacted by the time demands of the course and the perceived difficulty of the course work (8). The power-distance nature of undergraduate students’ interactions and relationships with clinical supervisors, is also perceived to cause stress (9). Within a clinical setting, undergraduate students also experience stress due to increased working hours, insufficient peer support and financial problems (10, 11). The clinical assessments are also source of anxiety, to optometry students, although the anxiety does not correlate with the clinical performance in the assessment (12).

About a third of qualified optometrists surveyed in Australia do report symptoms of anxiety and depression, with the study completed prior to COVID-19. These symptoms were more than threefold prevalent in practitioners under 30 years old (13). In job satisfaction interviews with working optometrists perceived their work to be stressful. Sources of stress were related to clinical decision-making and workload demands (14).

Optometry degrees have been taught face-to-face for many years. The pandemic caused disruption in normal academic scheduling, which has impacted the way students have been engaging with academic material. The 2020-2021 academic year saw optometry schools (and universities in general) forced into trying new types of learning. This meant various online modes of content for theory classes and smaller group learning for practical and clinical work. At present there is one study that investigated the wellbeing of optometry students, under normal teaching circumstances (2). The impact of the change of the educational interface and the ongoing pandemic on the wellbeing of optometry students is of value.

### Aim

From previous studies, predominantly into medical student experiencing stress and anxiety from their course, it is evident that there is an array of factors that may influence students’ wellbeing. There is a need to identify the exact factors influencing undergraduate optometry students’ wellbeing. Optometry students’ wellbeing, and more specifically stress and anxiety factors have not been investigated and were of particular interest to the researchers during the initial period of the pandemic. There is a gap in the knowledge on the status of optometry students’ wellbeing and what factors contribute to the wellbeing in optometry students.

The overarching aim of the study was to investigate wellbeing of optometry students. The study had two research questions:

I. What is the status of optometry students’ wellbeing?
II. Which factors are perceived to influence optometry students’ wellbeing?

This study took a quantitative approach with qualitative aspects, which was an observational cross-sectional descriptive study design.

### Sample

The target population (n=111) of this study was first to fourth (final) year optometry students registered at a South African campus in the year 2020. Students ages ranged from 17 to 32 years old. Inclusion criteria included students who were studying optometry and were willing to participate in the study. Though every effort was made to have the whole cohort contacted, due to the study being undertaken online, some students may have unintentionally been unable to take part, due to data access constraints.

## Ethical aspects

Researchers implemented precautions to ensure that the study was done in an ethically approved manner. Permission for study was obtained from the Health Sciences Research and Ethics Committee at the university where the data was collected. The participants gave prior consent and completed the questionnaire anonymously. The questionnaires that were used were only for screening purposes and did not diagnose participants with anxiety or depression. The contact details of the student wellbeing centre and the contact number of a qualified psychologist were provided to the participants on the information supplied before participant’s started the online questionnaire. Each participant was asked to supply their age and year of study, and no further demographic data. The information gathered was only accessed by the researchers and confidentiality was maintained throughout (11).

## Methodology

### Instrument

The instrument used in the study was a questionnaire comprising of two standardised sets of forced choice type questions and two open-ended questions. The two standardised tests were the Zung self-rating anxiety scale (SAS) and the Zung self-rating depression scale (SDS) (15, 16). These are standardised tests known to have been used in health profession students (6) and accepted as a valid method to assess an emotional response.

Open-ended questions aimed to get each participant’s own view of factors that influence their overall wellbeing and were asked to rank the factors from number 1 to 5. Number 1 representing the factor that is perceived as influencing their wellbeing as a student the most.

After the completion of the questionnaires, the open-ended questions data was thematically coded for analysis by the researchers.

### Pilot study

The online questionnaire was distributed to five voluntary fourth year optometry students, who determined the ease of understanding of the questions and determined the time it took to complete the questionnaire. The data from this pilot was included, as no changes were made to the open-ended questions and the Zung SAS and SDS are standardised questionnaires.

### Data collection process

Data collection took place in the first semester, during lockdown from April to May 2020, after ethical approval was granted. Due to students being at home, the method of contact was electronic. Students received an email with the link to the questionnaire along with an information sheet. Students could still choose not to participate by not completing the questionnaire once received. If students chose to participate, they filled in the questionnaire online and submitted it.

## Analysis of data

Analysis of the standardised components of the questionnaire was completed according to the scoring scales of the relevant questionnaires. The Zung SAS component of the questionnaire asked participants to indicate the factors of anxiety by marking the criteria most relevant to themselves, as follows; little of the time (=1), some of the time (=2), a good part of the time (=3) and most of the time (=4).

The Zung SAS overall scores ranged from 20-80. The participants select the appropriate score and the scores for all the questions are totalled. In the second component, the Zung SDS for each factor the numbers underneath the criteria are different, participants circled the number below the criteria most relevant to them. Zung SDS overall score ranged from 20-80. It was transformed to a SDS index score (17) with multiplying the raw score by 1.25, with a range of 25-100. The interpretation of the total of the raw scores indicates the level of anxiety and depression as seen in Table 1.

**Table 1:** Cut off values for Zung self-rating anxiety scale (16) and Zung self-rating depression scale (17)

| **Zung self-rating anxiety scale (SAS)** |  | **Zung self-rating depression scale (SDS)** |  |  |
| --- | --- | --- | --- | --- |
| **Interpretation:** | **Raw score:** | **Interpretation:** | **Raw score (20 – 80)** | **SDS Index (25 – 100)** |
| Normal range | 20-44 | Normal range | 20 – 39 | 25 – 49 |
| Mild to Moderate anxiety | 45-59 | Mild depression | 40 – 47 | 50 – 59 |
| Marked to severe anxiety | 60-74 | Moderate or marked depression | 48 – 55 | 60 – 69 |
| Extreme anxiety | 75-80 | Severe depression | 56 – 80 | 70 – 100 |

This scale has been determined to be a good discriminator between depressed and nondepressed individuals (18). With this the researchers determined what level of anxiety and depression the participant experienced according to the questionnaires’ scoring scale.

The open-ended questions underwent thematic analysis by frequency counts, where words or sentences were used as codes. The thematic analysis ustilised the steps described by Braun (19), was performed iteratively by all researchers, to ensure that the analysis was a true reflection of the voice of the participants. Four rotations of coding were undertaken, and themes were finalised by consensus. The frequency of statements related to the themes was also determined through word clouds, as an independent verification of the themes, in the absence of an external co-coder.

## Results

The response rate was 78.38-% (sample n=87; population n=111), comprising of first year (n= 31; 93.94%), second year (n=21; 65.63%), third year (n=18; 81.82%) and fourth year students (n=17; 70.83%).

### Standardised components of the questionnaire

Table 2 shows the results of the SAS and SDS. The majority (87.10%) of first year and all (100.00%) second year students were identified to have normal level anxiety symptoms. Moderate anxiety symptoms were present in a third (33.33%) of third year students and nearly a quarter (23.53%) of fourth year students. No year group presented with severe or extreme anxiety symptoms.

**Table 2:** Anxiety and Depressions Scales in undergraduate optometry students

| Student group | SAS |  |  |  | SDS |  |  |  |
| --- | --- | --- | --- | --- | --- | --- | --- | --- |
|  | Normal | Moderate | Severe | Extreme | Normal | Mild | Moderate | Severe |
| First Year | 87.10% | 12.90% | 0.00% | 0.00% | 51.61% | 32.26% | 9.68% | 6.45% |
| Second Year | 100.00% | 0.00% | 0.00% | 0.00% | 80.95% | 4.76% | 14.29% | 0.00% |
| Third Year | 66.67% | 33.33% | 0.00% | 0.00% | 50.00% | 11.11% | 22.22% | 16.67% |
| Fourth Year | 76.47% | 23.53% | 0.00% | 0.00% | 52.94% | 35.29% | 0.00% | 11.76% |

Depression symptoms were within normal range in the majority of the four groups of participants: 51.61%, 80.95%, 50.00% and 52.94% of the first to fourth year students respectively. It is noteworthy that severe depression symptoms were present in the first year (6.45%), third year (16.67%) and fourth year (11.76%) participants.

The one-way ANOVA test (p=0.005) indicated a statistically significant difference between the year groups’ results. The Turkey-Kramer ad hoc tests determined statistically significant differences between the year groups, as shown in Table 3 below.

**Table 3:**
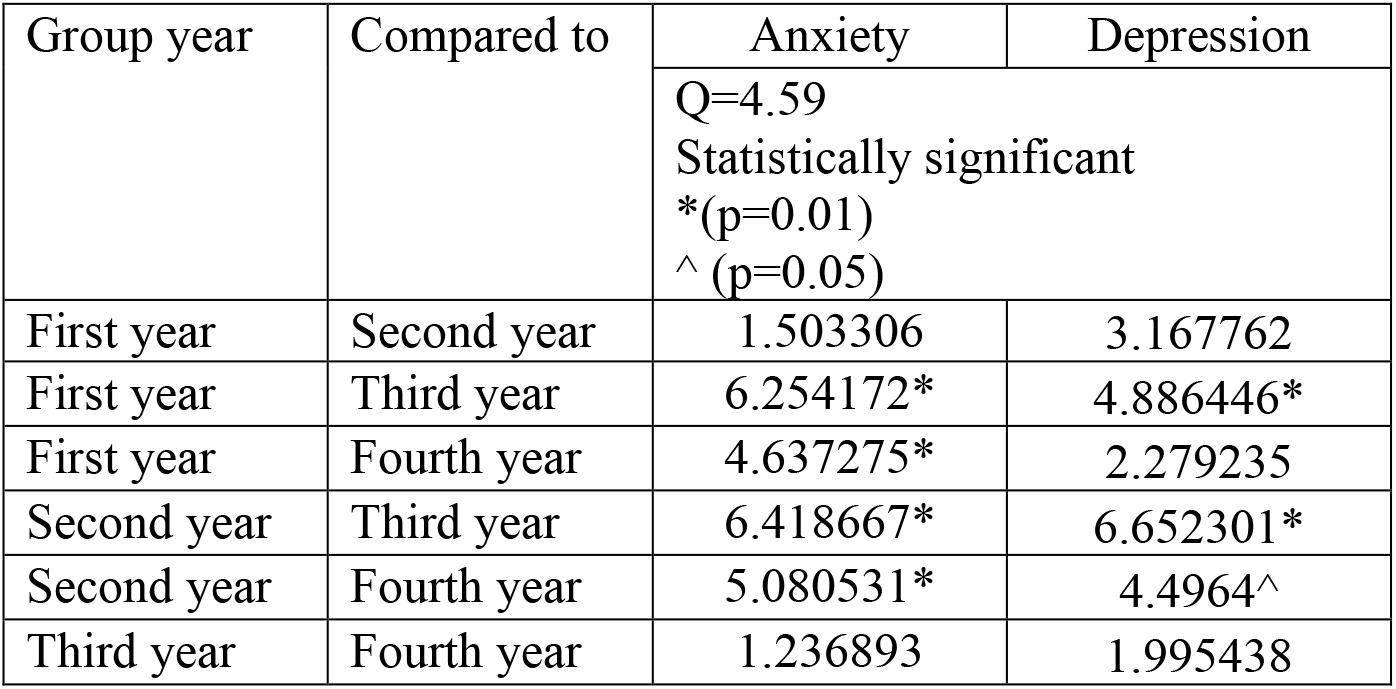
Turkey-Kramer results for all year group comparisons for Anxiety and Depression.

First and third year comparisons, and second and third year comparisons indicated a statistically significant difference (p=0.01) for both anxiety and depression scores. Whereas first and fourth year showed statistically significant difference (p=0.01) in their anxiety scores.

### Open-ended questions

The questionnaire ended with open-ended questions. Firstly, participants were asked to: *“Rank five general factors from 1 to 5 that has an influence on your wellbeing as a student. With 1 being the biggest influence and 5 being the smallest influence*.*”*

The responses were then grouped together to form the categories, and thereafter themes. After thematic coding the following themes emerged as the most important general factors influencing Optometry students’ wellbeing, provided in Table 4. Through frequencies of codes within the themes, the combined ranking for all year groups was established. The number appearing in brackets alongside the theme represents the number of statements coded within that theme.

**Table 4:**
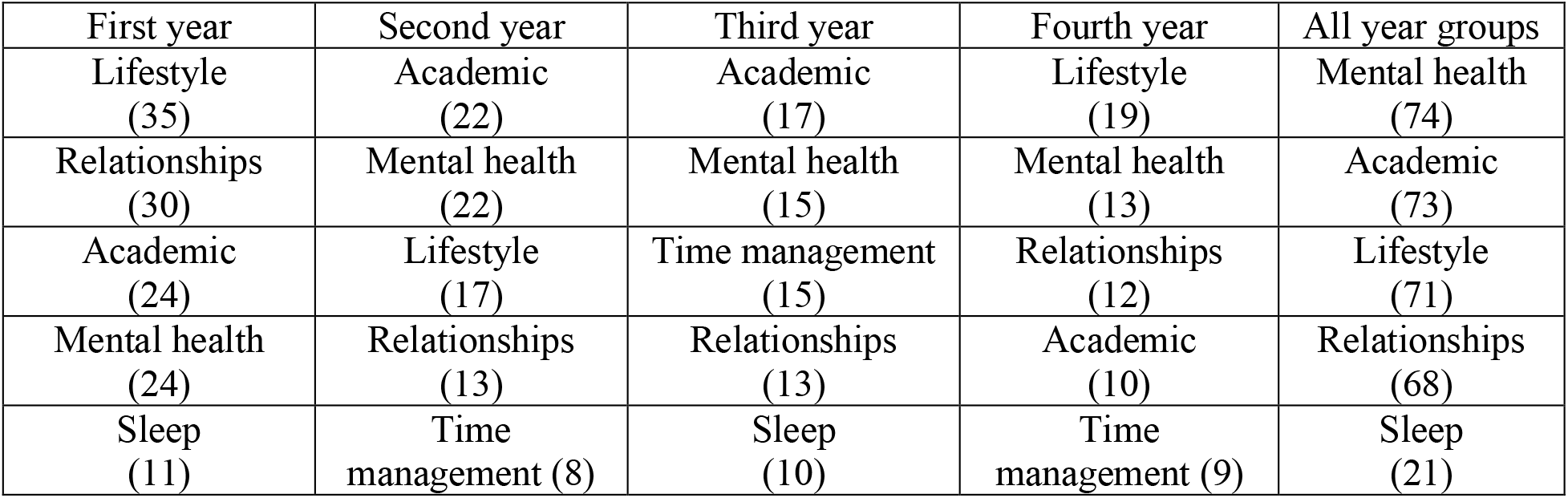
Ranking of general influencing factors on optometry students’ wellbeing

Mental Health statements included sub-themes of the participants perceived mental state, their religion and support networks, acts of self-care and sources of motivation. Academic statements included sub-themes of online learning, writing of tests, academic workload, the practical component of the course as well as the clinical rotations. Sub-themes in Lifestyle encompassed exercise, hobbies, finances, nutrition, hygiene and the use of alcohol and drugs. Relationships were made up of sub-themes of friends, family, social interactions and their partners. Many of the statements were one-word answers, such as sleep, tests, exercise and were coded as such.

The second open-ended question also asked students to list the most important factors impacting their wellbeing, with respect to COVID-19. The statements in response to this question were mostly full sentences; participants had a lot more of a voice with greater detail in this question. These statements provided by the participants speak to a more poignant reaction: “*losing out on life*” and “*will I be safe back at campus/clinic*”.

The top five factors that emerged from thematic analysis with respect to COVID-19 on optometry students’ wellbeing are presented below in Table 5. As in the previous open-ended question, the combined ranking for all year groups was determined by the frequencies of the codes within the theme. The value in brackets beside the theme indicates the number of statements coded within that theme.

**Table 5:** Ranking of COVID-19 specific influencing factors on optometry students’ wellbeing

| First year | Second year | Third year | Final year | All year groups |
| --- | --- | --- | --- | --- |
| Academic<br>(44) | Academic<br>(36) | Academic<br>(22) | Academic<br>(24) | Academic<br>(124) |
| Uncertainty<br>(17) | Uncertainty<br>(21) | Uncertainty<br>(17) | Physical Health<br>(21) | Uncertainty<br>(69) |
| Relationships<br>(15) | Physical Health<br>(11) | Change<br>(8) | Graduating<br>(16) | Physical Health<br>(47) |
| Mental health<br>(11) | Mental health<br>(9) | Relationships<br>(6) | Uncertainty<br>(14) | Relationships<br>(27) |
| Physical Health<br>(10) | Relationships<br>(6) | Physical Health<br>(5) | Change<br>(8) | Mental Health<br>(20) |

New sub-themes in this question pertaining to academics included studying methods and working independently, online learning, contact with lecturers and types of assessments. Uncertainty comprised of sub-themes of the future circumstances and safety. Physical health sub-themes pertained to fear of oneself or a family member contracting the COVID-19 virus, of being ill from the virus and death. Relationships included social distancing as a new addition that impacted their relationships with others. New sub-themes to Mental Health included conspiracy theories and fake news creating anxiety among participants.

Verbatim statements provided below give an overview of the severity of emotions elicited from the open-ended questions. There were concerns about academic performance impacted by the online method of teaching: *“Covid-19 affects the way we are taught new material, which is not always as ideal as face-to-face classes. This can make a concept more difficult to grasp, which can lead to increased stress levels*.*”, “Stress about failing”, “Feels like I am getting nowhere-worried about when we start writing paper tests again”*. First years did feel a lack of acclimation to the university life due to *“Lack of integration/adaptation to university life as a first year (how to prepare for exams/tests etc)”*.

Clinical rotations were a cause of distress for the students: “*Stress about clinic, seeing patients and disappointing supervisors”, “Struggling to sleep, because of stress for clinic the next day, tests and practical assessments”, “I am concerned about whether or not we will be able to catch up on all the clinic practice we missed while being at home*.*”*

Comments regarding the pandemic: “*Future of academic year as a result of the loss of time and the unstable conditions within the country and school “, “What happens with our practicals and clinic hours if this pandemic lasts until next year?”, “not being able to go to clinic for our practicals as we need them in order to complete our degrees (sic)”, “The virus has caused a lot of uncertainty and definitely contributed to a lot more stress on my shoulders”*.

Fear of being in the clinic: *“That my health would be at risk, and I contract the virus from a patient”, “…afraid to return to clinic and risk infection. I’ve been having nightmares about this …for the last few nights”*.

Personal circumstances: “*Whether my mother will lose her job”, “feel like a parent most of the time because I have to care for the household and my little brother most of the time…”*. Personal homelife conditions were also of concern, *“Timing of online tests, loadshedding, bad signal or no Wi-Fi…”, “domestic violence”* and *“drug abuse”*. Personal reflections, *“COVID 19 has taught me that I need to let go of the things I cannot control”; “I stress (about) a lot about things, yet I don’t get up and try to improve them”*.

The unusual circumstances during lockdown caused pause: *“Abuse of state/police/corporate authority”, “Scared the police will lock me up if my permit is not right”, “I worry if things will ever be normal again”, “…I would like it to end now”*.

## Discussion

It is pleasing to see that most participant cohorts did not self-identify with symptoms of anxiety. It is noteworthy that the third- and fourth year students perceive a moderate level of anxiety symptoms. These findings conform to the trend of higher year groups experiencing more anxiety, as found in medical students (3). This is reiterated in the statistically significant differences found between the first year and the third year as well as fourth year groups’ anxiety scores; and the first year and third year depression scores. The third year course is the heaviest workload in terms of course work with 11 theory modules and six practical components. There are fewer theory modules in the fourth year of the course. This may account for lower anxiety in the fourth years than in the third years, also as to why there is no statistically significant differences in the comparison of their perceived levels of anxiety and depression.

This study presents some similarity to a sample of practising optometrists (n=489) where less than half of the participants were found to have anxiety: mild (9.00%), moderate (18.40%), severe (5.70%) and extreme (7.00%)(13). Practising optometrists were found to have a higher level of depression when compared with the general public (13), with a study finding (n=489) 7.40% were displaying symptoms of extremely severe depression. The instruments used in this study differ from those used in practising optometrists, preventing a definitive comparison.

There was a wider range of perceived symptoms of depression, than anxiety. With more than half of each year group in the “normal” band of depression symptoms, this is similar to Maharaj (7). However, contrasts with results in medical students (6) which combined all scores above 50 in the Zung SDS to be indicative of depression. Using this cut off for depression, Basnet et al found 36.74% of first year medical students were depressed, compared to 48.39% optometry students in this study; and 22.22% of third year medical students (6), compared to 50.00% optometry students in this study, were depressed.

Hoying et al (2020) investigated health beliefs and behaviours in first year graduate health sciences students from seven health professions colleges, of which Optometry was one (n=3) (20). The Optometry students in this study did not present with any symptoms of anxiety and only one of the three students reported mild depression. With such a small sample of optometry students, it is difficult to draw a comparison to this study.

In a systematic review on medical students’ psychological distress, in the United States of America and Canada, it was identified that there is a high prevalence of depression and anxiety, compared with similar age range populations that are not healthcare students (3), as is the finding with practising optometrists (13). Sabih’s (2013) investigation on physiotherapy students found 40% of participants perceived mild stress and 6% reported experiencing high amounts of stress (21). It is evident from these findings that anxiety, depression and stress is present in other healthcare students, other than medical students. The findings in this study may point to perceived symptoms of both anxiety and depression in South African optometry students, although this cohort only represents one of the four Optometry Schools in the country.

### Open ended questions

COVID-19 has had a considerable impact on the academic experience of these participants, particularly due to online learning methods, a lack of practical and clinical time, greater requirement of independent study, and the resources to study. Blended learning is likely to continue and going forward more reliance of online learning is expected. This requires that students of the future learning these computer-based skills and being more accountable for self-directed learning. Due to the online nature of learning during COVID-19, students from less privileged households experienced more obstacles to academic success.

Fear and anxiety from the levels of uncertainty were expressed, both of which were seen as a negative impact on wellbeing. Future concerns have been noted in studies predating the pandemic (6). As COVID-19 is still a prevailing condition, it appears that students (and society as a whole) will need to be more accepting of change and learn to be more adaptable due to the ongoing climate of uncertainty.

The Relationships theme with sub-themes of family, social interaction and friends, it was friends that was the most frequently mentioned. Relationships obtained the highest ranking (second place) in the top five factors in the first year participants. During lockdown, this may have been a largely influencing factor, as students were living at home and spending the most time of with their family. A small number of students chose to remain in private student housing and were concerned about when they would be allowed to travel home.

Our findings on finance being a factor in general wellbeing, as well as that of concern for physical health, aligns with the study by Maharaj (7) and a review by Dyrbye (3). Students who were on bursaries paid a large portion of their funding to accommodation that was not being used, as students were unable to stay on campus. In addition, these students were unable to visit student finance offices with queries, as these services were closed during the lockdown. Actual job losses and the loss of income to students’ families during the lockdown were not directly mentioned, however this was inferred through comments of financial strain. The cause of this financial strain is thus likely to be different from these studies.

Sleep deprivation has been noted as a factor in medical students’ wellbeing (3). In a normal academic year, with many academic commitments, students are forced to be physically present in classrooms. The impact of needing sleep and not getting to rest was a frequently reported general factor perceived to influence wellbeing by both first- and third year students in this cohort. The maintenance of good sleeping-working balance (22) was no longer mandated by having to be present in face-to-face classrooms but fell to the student to manage independently. Students mentioned being awake all night and sleeping until midday, this routine caused them to skip meals and regular exercise.

The influence of COVID-19 on wellbeing showed a prioritisation of both physical and mental health, with physical health ranked in the top five factors in each year group. The concern of the higher year groups regarding their health and safety was mainly due the anticipated exposure to the virus during their clinical sessions. This concern extended to students showing concern for putting family members and house-share members at risk when they returned to clinics. In addition, the clinics were closed for an undetermined number of weeks, losing precious clinical time to make the required patient numbers, of which students were acutely aware. Furthermore, due to the limited numbers of patients being seen in the clinical sessions upon returning, this added to the anxiety and pressure to meet the required patient numbers.

Since it is apparent that students do experience some levels of anxiety as they progress through the optometry program, methods to assuage this should be considered. It is important that young adults learn how to develop a healthy lifestyle, this can be achieved through health promotion (22). Medical students who completed an elective course in mindful-based stress-reduction, using insight, relaxation and fostering concentration, seemed to have reduced the adverse effects of stress in mental health (3).

### Limitations

It is plausible that an increased response rate could have been achieved if the intended face-to-face distribution methods of data collection were possible. Findings in this study could be overestimated due to self-selection bias, where those who were experiencing lowered wellbeing were more likely to participate. Equally, students with diagnosed mental health problems may have chosen not to participate in this study for fear of being identified and feeling shame regarding their answers, even though the questionnaire was anonymous and confidential. Similarly, without a baseline level for anxiety and depression prior to COVID-19, it is plausible that these results represent a higher level of symptoms in this cohort, than would have been presented in any other year.

Methodologically the study could have been enhanced by having a third-party co-coder during the analysis to broaden the coding and to prevent bias. This was mitigated through the consensus method and the generation of word clouds, however, could be considered an enhancement for future studies.

## Recommendations

It is recommended that the study be repeated in the future, as scarce studies into optometry students’ wellbeing were found for comparison. Future studies could lead to the establishment of the baseline wellbeing of optometry students, not only in South Africa. It would provide some contextual baseline to repeat the study in a non-pandemic year and compare intervarsity findings among the four South African optometry departments.

An ongoing investigation could be embedded into the induction proceedings of optometry departments, to aid monitoring. In addition, measures to improve the wellbeing of optometry students could be implemented through early identification. At present the main method of management is the referral to the wellbeing clinic on campus. Anecdotal evidence from students, suggests this is neither adequate, nor effective. Moreover, responses from students in this study argue for more formal engagement of the department to support student wellbeing. Next steps should identify practical interventions to promote wellbeing and to prevent and assist optometry students with mental health symptoms and conditions.

## Conclusion

It with concern that can be concluded that nearly half of the participant optometry students self-identify with depression symptoms based on the SDS criteria. A moderate number of participant optometry students identify challenges that influence their optimal wellbeing. The dearth of any qualitative research of optometry students’ wellbeing prevents further comparison. The impact that COVID-19 has had on the wellbeing of optometry students is evident, and the contribution to the findings in this study, undertaken during lockdown circumstances, cannot not be taken as a reliable baseline. Further research into the wellbeing of optometry students is needed to verify the prevalence the concerns identified in this study.

## Data Availability

Data for this manuscript is available upon reasonable request, as regulated by the approving ethical body for this study.

## Acknowledgements

We thank the participants for taking the time to submit the questionnaire whilst in lockdown.

